# A SARS-CoV-2 Reference Standard Quantified by Multi-digital PCR Platforms for Quality Assessment of Molecular Tests

**DOI:** 10.1101/2020.09.14.20193904

**Authors:** Haiwei Zhou, Donglai Liu, Liang Ma, Tingting Ma, Tingying Xu, Lili Ren, Liang Li, Sihong Xu

## Abstract

SARS-CoV-2 is the seventh coronavirus known to infect humans and has caused an emerging and rapidly evolving global pandemic (COVID-19) with significant morbidity and mortality. To meet the urgent and massive demand for the screening and diagnosis of infected individuals, many in vitro diagnostic assays using nucleic acid tests (NATs) have been urgently authorized by regulators worldwide. The limit of detection (LoD) is a crucial feature for a diagnostic assay to detect SARS-CoV-2 in clinical samples, and a reference standard with a well-characterized concentration or titer is of the utmost importance for LoD studies. Although several reference standards of plasmids or synthetic RNA carrying specific genomic regions of SARS-CoV-2 have already been announced, a reference standard for inactivated virus particles with accurate concentration is still needed to evaluate the complete procedure including nucleic acid extraction and to accommodate customized primer-probe sets targeting different genome sequences. Here, we performed a collaborative study to estimate the NAT-detectable units as viral genomic equivalent quantity (GEQ) of an inactivated whole-virus SARS-CoV-2 reference standard candidate using digital PCR (dPCR) on multiple commercialized platforms. The median of the quantification results (4.6×10^5^ ± 6.5×10^4^ GEQ/mL) was treated as the consensus true value of GEQ of virus particles in the reference standard. This reference standard was then used to challenge the LoDs of six officially approved diagnostic assays. Our study demonstrates that an inactivated whole virus quantified by dPCR can serve as a reference standard and provides a unified solution for assay development, quality control, and regulatory surveillance.

## Introduction

COVID-19 is the disease associated with SARS-CoV-2 infection and is an emerging and rapidly evolving worldwide pandemic^
1,2
^. This pandemic has affected more than 188 countries or regions with more than 18,575,326 confirmed cases and 701,754 confirmed deaths as of August 6, 2020^3^. Person-to-person transmission from asymptomatic individuals has been reported and can exacerbate the spread of the pandemic^4^. A major challenge faced by assay developers and health authorities is providing clinical diagnostic tests with a good limit of detection (LoD) and accuracy for the screening and diagnosing of those suspected of having COVID-19, even when the virus is present in minute quantities. Clinical diagnosis can identify infected individuals including asymptomatic people. This can inform patient care and disease management, suppress infection and transmission, and provide epidemiological and surveillance information. To meet the urgent and massive demand for diagnostic tests, the China National Medical Products Administration (NMPA; formerly, CFDA) and other health authorities have urgently authorized a large number of in vitro diagnostic assays based on nucleic acid tests (NATs)^5^. Although LoD is a part of the product specification, a direct comparison to assess and benchmark those assays under well-defined settings remains unexplored.

To determine the LoDs of diagnostic assays, various reference materials such as synthetic RNA transcripts, extracted genomic RNA, armored RNA, or inactivated virus particles with known concentrations or titers are commonly used. The use of various reference materials without consolidation leads to inaccurate and improper determination of LoDs of different diagnostic test kits—especially during an emergency such as COVID-19.

Virus particles are the ideal reference standard because they can evaluate the entire process, from nucleic acid extraction to results reporting. The reference standard must be appropriately quantified. Discrepancies in reference standards hinder attempts to understand assay performance by both assay developers and regulatory agencies because the same diagnostic assay can deliver very different performance metrics depending on which reference standard is used. Malfunctioning assays due to poor precision of reference standard will suffer issues such as low positive rates in the field, inconsistent test results, delays in clinical action, and the premature release of sick people.

Digital PCR (dPCR) is an emerging technology that achieves the absolute quantification of nucleic acid targets in a sample by partitioning analyte molecules into a large number of miniaturized reaction volumes, followed by the Poisson statistical analysis of the end-point binary results from each partition^
6–9
^. Due to its superior sensitivity, specificity, and absolute quantification ability, dPCR has been used for the diagnostic detection of SARS-CoV-2 as well as the quantification of reference standards—especially when the calibration material is not readily available^
10–13
^. For example, synthetic viral RNA, extracted genomic RNA, and inactivated virus particles offered by Biodefense and Emerging Infections Research Resources Repository (BEI resources) were quantified with a Bio-rad QX-200 Droplet Digital PCR System. However, quantification efforts that rely on one particular platform can deliver biased measurements due to technical, environmental, and biological factors. This systematic error, if not well characterized and understood, can lead to adverse effects with different NATs. We aim to overcome this challenge through recruiting several distinct dPCR platforms in our study.

This work reports a multi-platform dPCR quantification of a SARS-CoV-2 reference standard candidate via inactivated whole-virus particles in collaboration with nine independent participants equipped with six dPCR platforms. Several estimators were explored to determine the consensus true value of the concentration of virus particles in the reference standard. Finally, this reference standard was used to challenge the LoDs of six diagnostic assays approved by WHO and China NMPA.

## Methods and Experiments

### Preparation of reference standard candidate

A SARS-CoV-2 culture (BetaCoV/Wuhan/IPBCAMS-WH-01/2019, GenBank: MT019529.1) was provided by the Institute of Pathogen Biology, Chinese Academy of Medical Sciences & Peking Union Medical College. The strain was cultured with Vero cells (ATCC® CCL-81) at 37°C in a 5% carbon dioxide incubator. The cultured viruses were frozen and thawed once at −80°C; cytopathic effects (CPE) were observed in more than 60% of cells. The cultured viruses were inactivated by heating at 56°C for 1 hour (h). The collected viruses were then centrifuged to remove cell debris at 3000 g, 4°C for 10 min. The concentration of the supernatants was determined using a digital PCR detection kit (TargetingOne Corporation, China). The SARS-CoV-2 stocks were diluted to ∼ 1×10^6^ GEQ/mL in a universal buffer (10 mM PBS buffer pH 7. 5, 1% human serum albumin, 0.1% trehalose). Bulk preparations were aliquoted into 2 ml screw-cap tubes and stored at –80°C for inclusion in the collaborative study. This sample was provided to nine independent participants equipped with six different dPCR platforms, and the experiments were performed immediately upon sample arrival.

### Viral RNA extraction and One-step RT-dPCR

The same viral RNA extraction kit and protocol were adopted by all participants in the quantification of the reference standard candidate. Viral total RNA was extracted with the QIAamp Viral RNA Mini Kits (Qiagen, Hilden, Germany) in biosafety level-2 laboratories (BSL-2). A volume of 140 μL virus sample was used for RNA extraction, and the extracted RNA was eluted in 60 μL RNase-free water. The RNA extraction process was performed in duplicate for the quantification of reference standard candidate. To quantify the SARS-CoV-2 reference standard candidate using digital PCR method, three participants used a QX-200 system (Bio-rad, CA, USA), two participants used the TD-1 system (TargetingOne, Beijing, China), and the other four participants used the Naica system (Stilla Technologies, Villejuif, France), OsciDrop Flex system (Dawei Bio, Beijing, China), Starry 10K system (DAAN Gene, Guangzhou, China), and MicroDrop-100 system (Forevergen, Guangzhou, China).

### Estimation of measurement uncertainty

The concentration of virus particles for ORF1ab and N gene in the reference standard candidate was reported by the participants. The replicate measurements from extraction were combined to calculate the mean value of concentrations for each gene. The variance from extraction and dPCR replicates was calculated through a one-way ANOVA in R^14^.

The standard uncertainty of the measurement data was calculated by taking the square root of the sum of the variance divided by the number of replicates^15^. The relative standard uncertainty of the measurement data was obtained by dividing the standard uncertainty of the measurement data by the mean concentration for each gene. Finally, the relative standard uncertainty for the concentrations of ORF1ab and N genes was determined by combining the relative uncertainty of measurement data and the relative uncertainty of droplet volumes from instrument specification^
16,17
^.

### Estimation of the consensus true value of the concentration of virus particles

All candidate Key Comparison Reference Value (KCRV) estimators were calculated in R. The mean, median, and their associated uncertainties were calculated with native R functions. The DerSimonian-Laird (DSL) procedure and restricted maximum likelihood (REML) fits were calculated from the metrology package. The uncertainty-weighted mean was calculated from the glm function in the stats package. The Huber estimate 2 (H15) was calculated from the MASS package. The uncertainty-weighted Huber estimate was calculated with rlm function from the MASS package.

### Evaluation of LoD of commercial diagnostic assays

The quantified reference standard was serially diluted and applied to LoD probit regression analysis for six diagnostic assays. A total of at most 10 concentration levels were tested with multiple replicates per concentration according to the manufacturer’s instructions with an additional 10 replicates of viral transport medium as the blank control. Probit regression analysis of 95% hit rates was performed with SPSS 16.0 software (SPSS Inc, Chicago, IL).

## Results and discussion

### Study design for quantification of reference standard candidate

We adopted a multi-platform strategy to achieve accurate quantification of the SARS-CoV-2 reference standard candidate. We hypothesized that platforms and assays with different properties such as precision and accuracy of droplet volume, number of effective droplets, or assay threshold for distinguishing positive and negative reactions would introduce supplier-dependent systematic errors into the measurements. Therefore, we included a variety of dPCR platforms and service laboratories in this study. A reference standard candidate containing heat-inactivated virus particles was provided to nine participants equipped with six dPCR platforms including four new digital PCR platforms introduced by Chinese companies. The technical specifications and key features of the dPCR platforms are summarized in Table 1. These platforms operate through droplet microfluidic chips, microwells, and Droplet Array Production by Cross-interface Oscillation (DAPCO^TM^) technologies and generate droplets ranging from 10,000 to 100,000 with a claimed precision of droplet volume between 2.8% and 10%. Two to four fluorescence channels are available for hydrolysis of the fluorescence probe-based assays.

**Table 1.**
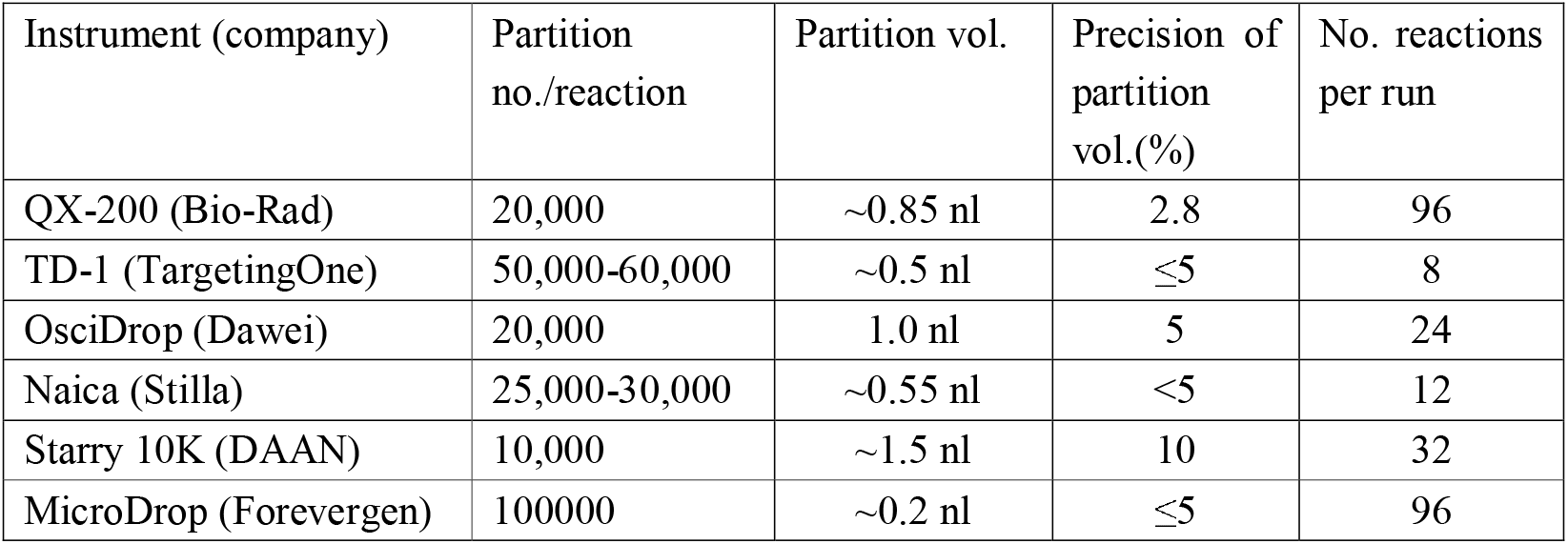
Technical specifications of dPCR platforms used in this study.

The study comprises two major steps: RNA extraction and one-step RT-dPCR. Virus particles were used as the starting material to ensure that the entire workflow is evaluated, and variations in each step contribute to the precision and accuracy of quantification. They thus provide a good estimate for real-world situations in clinical diagnostic settings. QIAamp Viral RNA Mini Kit is one of the kits recommended by the US CDC and was used by all the participants to extract total viral RNA. Due to logistical constraints, two replicate extractions were performed by each participant. All participants were required to follow the same RNA extraction protocol. A 5-step, 10-fold dilution series of the starting material was prepared, and each dilution was measured with an individual dPCR platform to determine which dilution falls into the linear digital quantification range to report an accurate quantification result. One-step RT-PCR reactions were performed to ensure consistency and minimize cross-contamination. Reverse transcription was initiated after droplet formation with gene-specific primers to synthesize cDNA that was used as PCR templates in the following hydrolysis probe-based assays. Each participant adopted their own experimental conditions including primer-probe sets, input amounts of RNA templates, RT-PCR supermix, cycling conditions, and dPCR platforms: These were optimized for the RT-PCR assay-dPCR platform combinations and were necessary to assess quantification results as practical dPCR assays at each laboratory.

The one-step RT-dPCR assays were performed in two formats: singleplex and duplex. Singleplex assays are straightforward to implement. Each data point is an independent measurement because the reaction is performed separately. Although multiplex assays may be influenced by primer/probe interactions or competition for resources, they facilitate simultaneous amplification of multiple target sequences in the same tube for the same sample. Singleplex dPCR assays targeting the ORF1ab and N regions were performed by five participants, and duplex dPCR assays targeting the ORF1ab and N regions simultaneously were performed by four participants. The quantification results were reported by the participants and analyzed to determine the consensus true value of the virus GEQs.

### Determination of the viral GEQ of the reference standard

We first visualized aggregate results from all participants and assays with a box plot, a violin plot (Figure 1A) and histogram (Figure 1B). Overall, the results showed a high degree of consistency on a coarse scale with means of 4.88×10^5^ GEQ/mL (CV = 43%) and 4.22×10^5^ GEQ/mL (CV = 44%) for the concentrations of ORF1ab and N gene, respectively. This agreement is remarkable in light of the differences in instruments, assays, experimental conditions, and operators between all nine participants. There is no calibration curve or internal control. This highlights the unique capability of dPCR for accurate and absolute quantification of nucleic acids. Technical duplicates from RNA extraction demonstrated high reproducibility within each participant. Considering the variation in droplet volume, the relative standard uncertainties of concentrations of ORF1ab and N genes in the sample for all suppliers showed decent precision and was less than 10% for most suppliers (Figure 1C).

**Figure 1.**
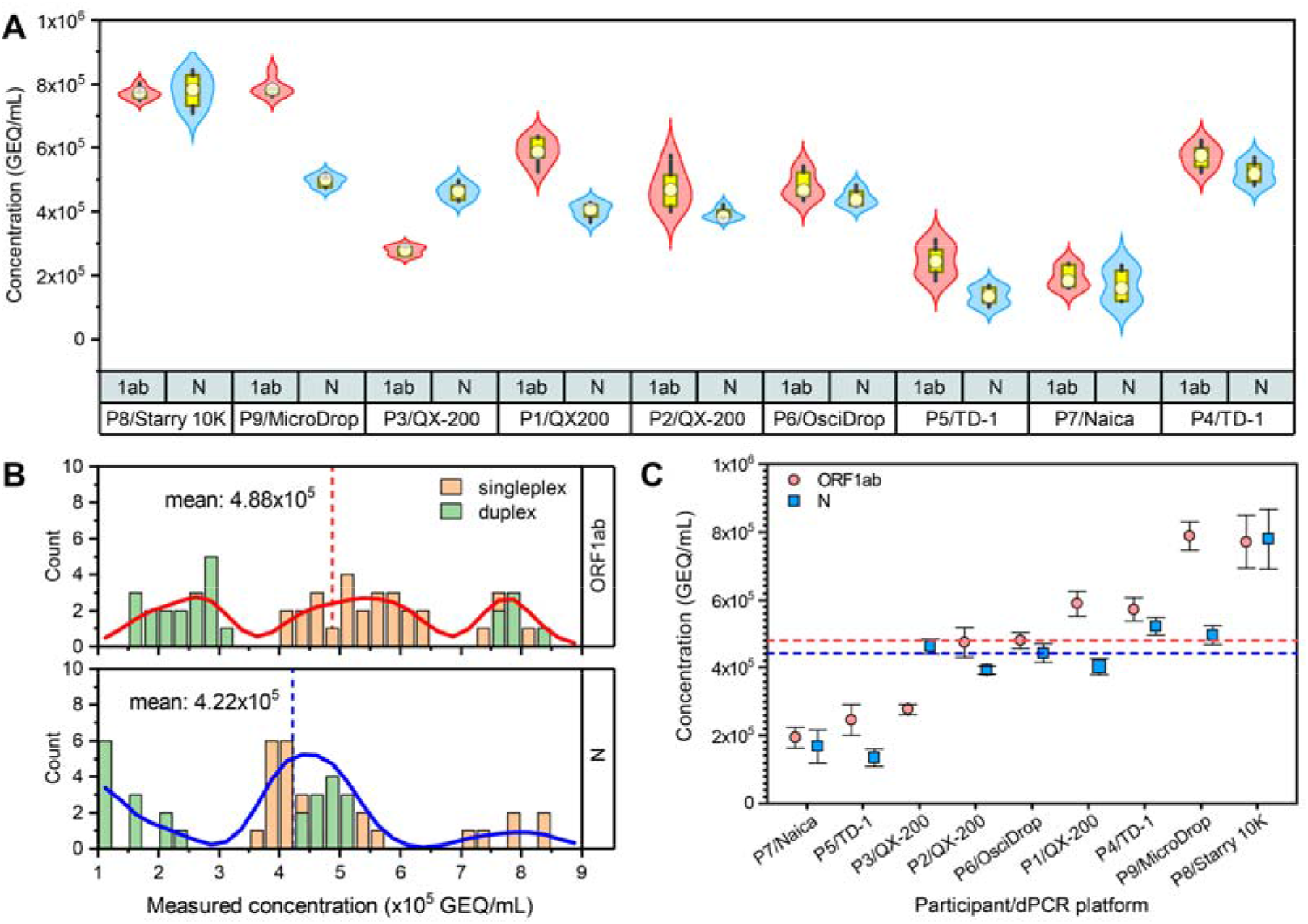
Overview of quantification results of ORF1ab and N gene of the reference standard candidate by nine participants using six dPCR platforms. (A) Box plots overlaid with a violin plot from three measurements of replicate RNA extraction (total of 6 measurements). Singleplex assays were performed using Starry 10K, TD-1 (by participant 4), OsciDrop, and QX-200 (by participants 1 and 2) platforms. Duplex assays were performed using Nacia, MicroDrop-100, TD-1 (by participant 5), and QX-200 (by participant 3) platforms. For each participant, the red box plot on the left shows the concentration for the ORF1ab gene, and the blue box plot on the right shows the concentration for the N gene. (B) Histogram of aggregate data sets (six measurements from individual participants for each gene). The wavy curves in the histograms represent smoothed density, and vertical dashed lines indicate median values. (C) Reported mean concentration with standard uncertainties by each participant taking droplet volume precision into consideration. Horizontal dashed lines represent the median values for the mean concentrations of ORF1ab (red) and N (blue) genes.

At fine scale, however, the variation of the measured number of copies of ORF1ab and N gene with different dPCR platforms is apparent. Histograms for both ORF1ab gene and N gene showed dispersal with a multi-modal distribution (Figure 1B). These data indicate heterogeneity within the data sets. The variation presumably stems from anticipated substantial differences in platforms, plexy, and assay design as well as the RNA extraction induced variation. In particular, we looked at the effect of assay plexy on the quantification outcome. The duplex assay gives a higher variation. The relative standard deviations (RSD) of measured mean concentrations with singleplex dPCR are 20.8% and 31.5% for ORF1ab and N regions; those with duplex dPCR are 73.5% and 60.3%, respectively. Our attempt with a t-test and two groups of data sets: singleplex (n = 4) and duplex (n = 5) on the mean concentration of each gene analyzed resulted in an insignificant difference (p>0.1). However, the effect of assay plexy on measured concentrations could not be rigorously determined due to the confounding factor of the dPCR platforms. Therefore, both singleplex and duplex data sets were included in the subsequent analysis.

### >Estimation of consensus true value of the concentration for virus particles

The KCRVs were estimated following the Consultative Committee for Amount of Substance (CCQM) guidance note CCQM13–22^18^. The entire data set was used to calculate the candidate KCRV estimators. The results are plotted in Figure 2. The candidate KCRV for the concentration of ORF1ab gene varied from 3.9×10^5^ to 4.9×10^5^ GEQ/mL. Those for the concentration of the N gene varied from 4.0×10^5^ to 4.4×10^5^ GEQ/mL. Most participants presented precise measurements with similar uncertainties. We considered seven estimators including estimators that do not leverage reported uncertainties in location estimation such as mean, median, and H15 estimates. Given the dispersion and heterogeneity within the data set, a median estimator was used for KCRV assignment because of its resistance to outliers. The median is 4.8×10^5^ GEQ/mL with a standard uncertainty of 1.3×10^5^ GEQ/mL for the concentration of ORF1ab gene, and 4.4×10^5^ GEQ/mL with a standard uncertainty of 3.4×10^4^ GEQ/mL for the concentration of N gene. We used the arithmetic mean of concentration of ORF1ab gene and N gene to calculate the concentration of virus particles. The standard uncertainty of the reference standard was obtained by combining of the standard uncertainties of ORF1ab and N genes. The concentration of virus particles is determined to be 4.6×10^5^ GEQ/mL with a standard uncertainty of 6.5×10^4^ GEQ/mL.

**Figure 2.**
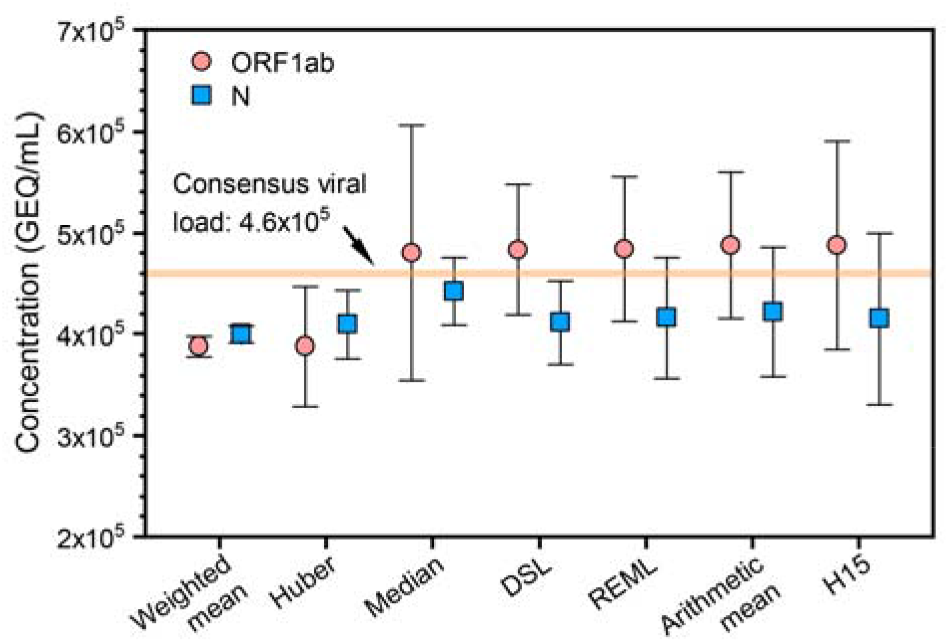
Calculation of Key Comparison Reference Values (KCRVs) and viral load estimated by the average of ORF1ab and N genes. A median estimator was chosen for KCRV assignment.

### Evaluation of LoD of the diagnostic assays

We applied the reference standard with consensus true value to evaluate the LoDs of six diagnostic assays approved by WHO or China NMPA (Figure 3). The tests cover one-step RT-PCR assays that require an extra step for nucleic acid extraction or release (Sansure, BGI, and Liferiver), fully automated NAT following the conventional workflow (Roche), and cartridge-based POCT devices for rapid “sample-in-answer-out” diagnostic tests (Cepheid and Ustar). According to the manufacturers’ instructions, the claimed LoDs of these diagnostic assays were obtained using reference materials such as armored RNA and clinical specimen RNA. Using the reference standard established in this work, we measured LoDs for all target genes of six diagnostic assays based on probit regression analysis of 95% hit rates (Fig. 3). Our study demonstrates that most diagnostic assays included in the study can meet or exceed the claimed sensitivity and are reliable for SARS-CoV-2 detection. All 12 claimed LoDs fall within the two-fold region of the measured LoDs. The results show that the reference standard is suitable for the assessment of various NATs and allows for comparable LoD studies of various detection methodologies. However, further validation using clinical samples is required to thoroughly evaluate the sensitivity and specificity of these diagnostic assays. For example, additional features such as reliability with poor quality and stability of viral RNA in the clinical sample, high specificity against background genetic material, and robustness to PCR inhibitors may further stratify the performance of diagnostic assays in clinical settings.

**Figure 3.**
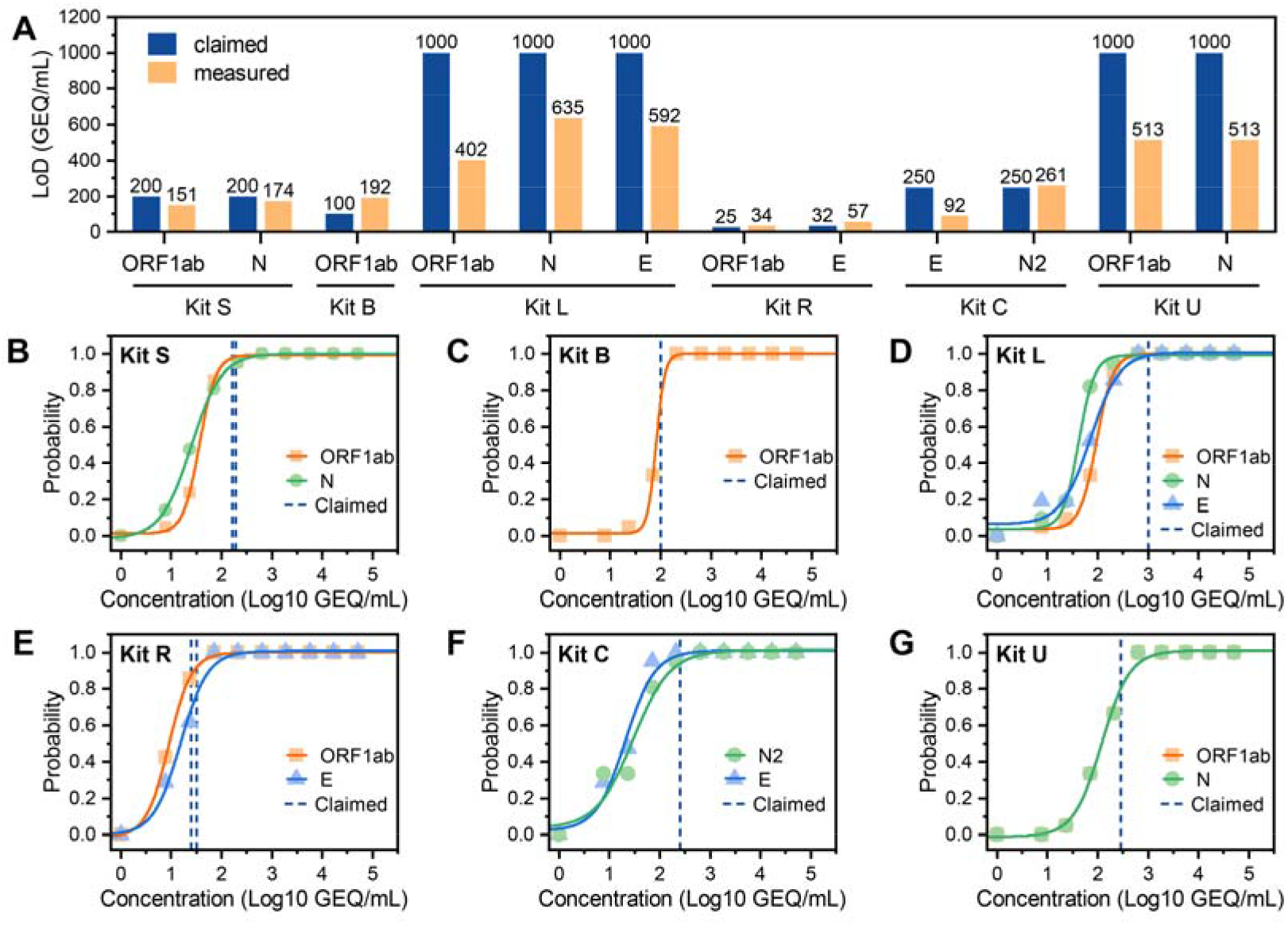
(A) Comparison of claimed LoDs of six NAT kits by the manufacturers with the measured LoDs using the reference standard candidate. (B-G) Probit regression analysis of six authorized diagnostic assays for SARS-CoV-2 molecular testing (SPSS). The probit (predicted proportion of positive replicates) versus the SARS-CoV-2 concentration was obtained by 21 replicates of 10 serial dilutions and an additional 10 replicates of a blank sample.

## Conclusion

This paper established a quantitative reference standard for inactivated SARS-CoV-2 whole-virus particles cultured from a representative isolate with a multi-platform dPCR approach. The Clinical and Laboratory Standards Institute (CLSI) recommends the use of WHO or relevant reference materials with accurate assigned values for the LoD determinations of diagnostic assays, especially for NATs of infectious diseases^19^. The reference standard established in this work was prepared using an inactivated whole-virus culture that was accurately measured using multi-digital PCR platforms. The standard can be used for LoD determinations during assay development as well as assessment. Thus, in comparison with existing reference materials such as synthetic RNA, genomic extraction, or pseudoviruses, the newly established reference standard is more suitable for the determination of the LoDs of various NAT assays regardless of assay platforms, extraction methods, and target genes.

We leveraged the unique capability of dPCR for the absolute quantification of nucleic acid samples and adopted a variety of dPCR technologies with distinct partitioning mechanisms and technical features to achieve the consensus true value of the concentration of virus particles through statistical analysis of the data set and judicious choice of estimators. The use of multiple platforms effectively corrects the system-level deviation from the true value due to the use of a single digital PCR platform^
20,21
^.

However, this collaborative and multi-platform emergency research also has some limitations. First, we found that single-plex dPCR provides lower inter-laboratory variation than multiplex dPCR and is better suited for the quantification of reference materials. However, the effect of the plexy of digital PCR for quantification was not adequately addressed due to limitations in the scope of the study as well as constraints from commercial suppliers. Second, the claimed droplet volume precision by manufacturers was used to estimate its contribution to measured uncertainty while variations in droplet volume were not experimentally determined and provided by the participants or the manufacturers. Third, the efficiency of nucleic extraction and operator-induced variability were not thoroughly assessed due to the difficulty of using an inter-laboratory study design to accommodate many replicates. Therefore, data sets with some participants may introduce random errors in addition to the variation of dPCR platforms. Future research should consider these limitations for better validation of the reference materials. Despite these constraints, our results were derived from the consensus of a large number of relatively uncorrelated dPCR platforms and service suppliers with distinct characteristics. The consensus quantification should outperform its individual constituents for accuracy and precision.

Overall, our findings showed how the quantification result can be influenced by platforms and assays, and a multi-platform collaborative effort is a promising approach for ensuring quality and accuracy in a timely manner. A reference standard using inactivated whole-virus particles ensures a thorough evaluation of the complete *in vitro* diagnostic package, including nucleic acid extraction, amplification, and detection. The results offer a unified solution for assay development, quality control requirements, and regulatory surveillance.

## Data Availability

Some or all data, models, or code generated or used during the study are available from the corresponding author by request. (List items).

## Acknowledgements

This work was financially supported by the National Science and Technology Major Project of China (2018ZX10102001) and the National Key Research and Development Program of China (2019YFA0904700).

